# Study on the usefulness of Direct Saliva sample Collection (DiSC) by polyester swab

**DOI:** 10.1101/2022.02.25.22271529

**Authors:** Kotaro Aoki, Mami Nagashima, Katsuhito Kashiwagi, Takashi Chiba, Kenji Sadamasu, Yoshikazu Ishii, Kazuhiro Tateda

## Abstract

Saliva sample can be self-collected and used in testing of SARS-CoV-2 nucleic acid amplification tests (NAATs) test in Japan. However, this may have difficulty collecting a proper specimen when collecting for the first time. We compared 2 collection methods, conventional methods and Direct Saliva Sample Collection method (DiSC) from 44 asymptomatic or symptomatic individuals who were in quarantine in Toho university hospital. RT-PCR by DiSC method showed about 70 % positive percent agreement compared to RT-PCR by conventional methods. In addition, comparing RT-PCR and TMA by DiSC method, TMA showed about 90 % positive percent agreement compared to RT-PCR. DiSC method is easy to perform by every person, does not have complicated restrictions/instructions and can be used in RT-PCR and TMA. This method allows for ease of saliva collection in certain patient populations.

---

Accurate pathogen detection by nucleic acid amplification tests (NAATs) begins with effective and consistent specimen collection methods. Especially for SARS-CoV-2 testing, it is important to collect samples safely and consistently for best results and protection of healthcare workers. Currently, nasopharyngeal swab (NP), nasal vestibular swab, and saliva have been used as specimens in SARS-CoV-2 NAATs [1-3]. These three specimen types have advantages and disadvantages in consistency and simplicity of collection. For example, although NP is more consistently collected, it must be collected by a medical worker, who may be exposed to risk of droplet infection by the spray from patients [4]. Saliva can be self-collected and a relatively homogeneous sample can be obtained. However, the person, who is not a medical worker and does not have specimen collection experience, may have difficulty collecting a proper specimen.

In this study, saliva was collected from 44 asymptomatic or symptomatic individuals who were in quarantine in Toho university hospital. Saliva was collected directly from mouth by simply placing an Aptima® Multitest swab between the gums and the inner cheek for approximately 30 seconds (Direct Saliva Sample Collection method; DiSC); paired saliva samples were collected from the same individuals using conventional methods [5]. The swab was suspended in Aptima^®^ Multitest swab collection tube (MLT; Hologic, U.S.A.). We compared the DiSC with conventional collection method by using N2 RT-PCR (140 μL from MLT was used for extraction for RT-PCR) (5). We also compared a transcription mediated amplification (TMA) NAAT, the Aptima^®^ SARS-CoV-2 run on the Panther system (Hologic, U.S.A.), with the RT-PCR for the sample set collected by DiSC (n=44). The limit of detection for the N2 region RT-PCR used in this study is 50 digital PCR copies / mL in starting sample using SARS-CoV-2 Molecular Q Panel (RANDOX Laboratories, U.K.) and QIAamp Viral RNA mini kit (QIAGEN, Germany), TaqMan Fast Virus 1-Step Master Mix and QuantStudio^®^5 (ThermoFisher Scientific, U.S.A.) in our laboratory. The study design was approved from the Ethics Committee of Toho University School of Medicine (A20116_A20028_A20020_A20014_19099). The results indicate that while the DiSC method may have somewhat decreased sensitivity as compared to the conventional method, it still detected approximately 70% of positive samples (Table 1). Of the 5 DiSC specimens positive only by TMA, three had paired conventional collection saliva specimens that were positive by RT-PCR (Ct values 30-34) (Supp. Table 1). Our results also show that TMA detects more positives than the SARS-CoV-2 N2 RT-PCR for saliva collected by DiSC. World Health Organization document stated that it may be desirable to use a nasal swab sample rather than a saliva sample because the definitive diagnosis of SARS-CoV-2 requires high sensitivity and specificity, but screening tests to confirm cases with suspicious COVID-19 and its history of infection do not require high sensitivity [6]. DiSC is easy to perform by every person, does not have complicated restrictions/instructions (e.g., eating/drinking, time of day for collection, time to processing, etc), and may allow for ease of saliva collection in certain patient populations, particularly children, the elderly, and patients with collagen disease, while still maintaining acceptable sensitivity.

**Table 1.**
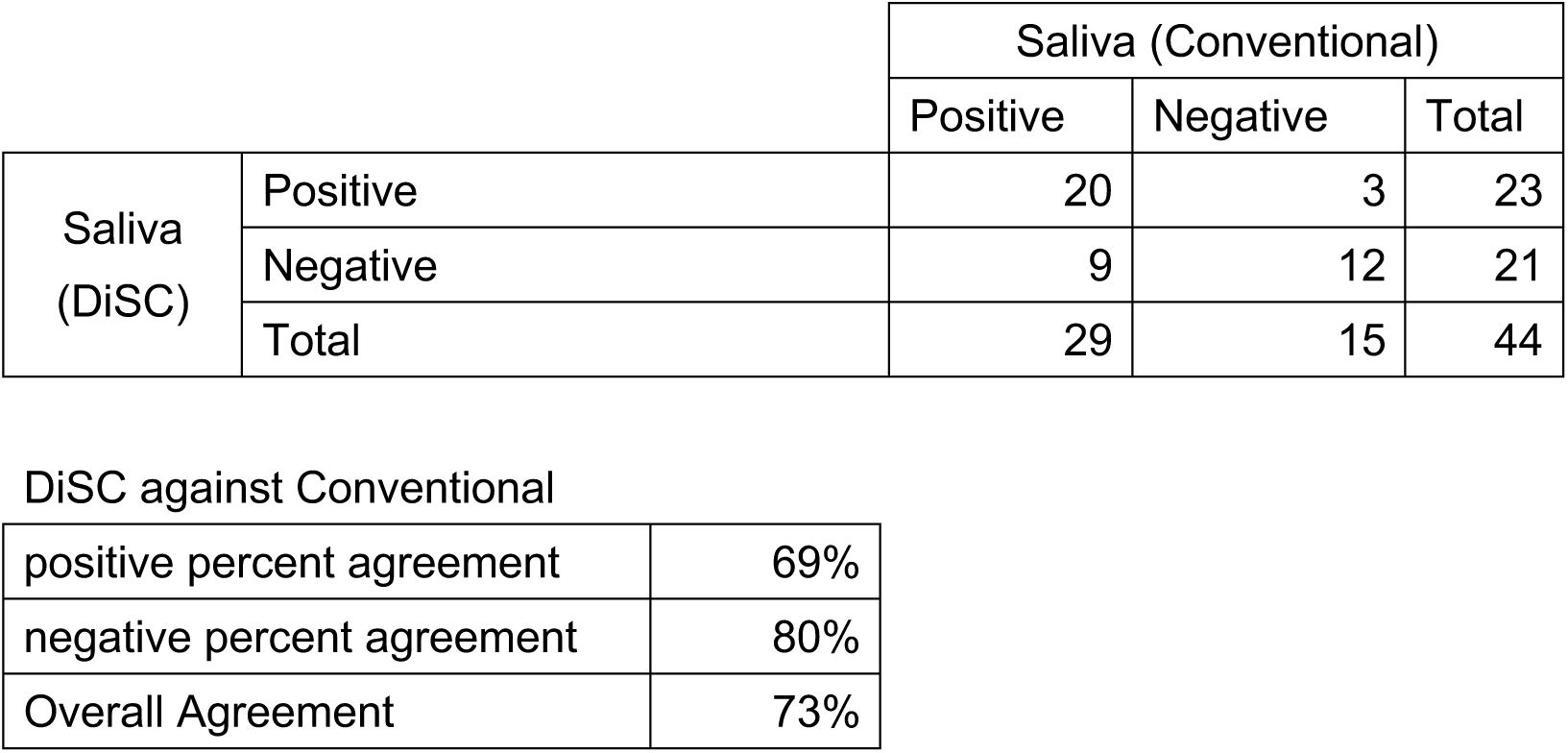
Evaluation results of the Direct Saliva Sample Collection method (DiSC) and the conventional collection method by RT-PCR.

|  |  | Saliva (Conventional) |  |  |
| --- | --- | --- | --- | --- |
|  |  | Positive | Negative | Total |
| Saliva<br>(DiSC) | Positive | 20 | 3 | 23 |
|  | Negative | 9 | 12 | 21 |
|  | Total | 29 | 15 | 44 |

|  |  |
| --- | --- |
| DiSC against Conventional |  |
| positive percent agreement | 69% |
| negative percent agreement | 80% |
| Overall Agreement | 73% |

**Table 2.**
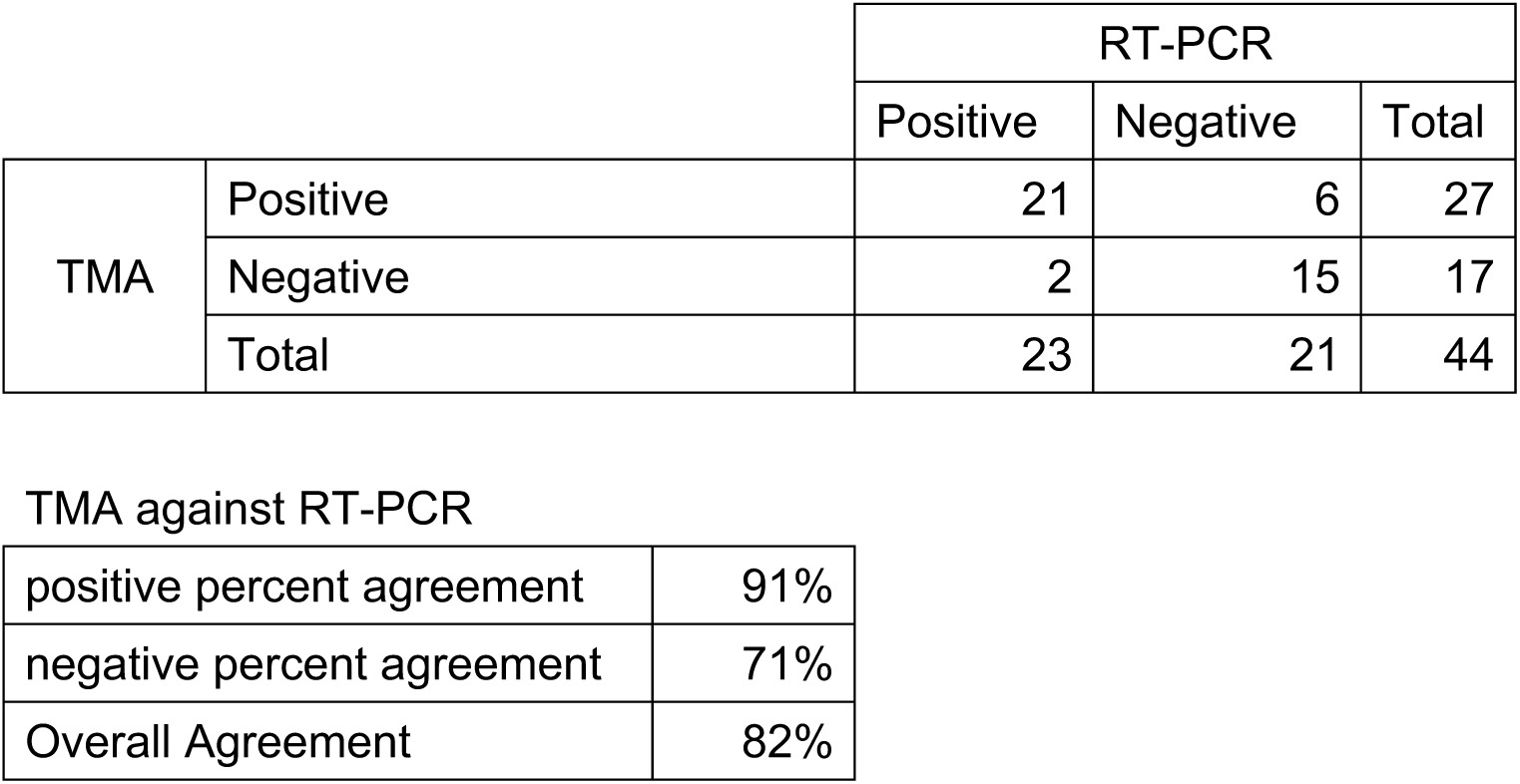
Evaluation results of the TMA method and RT-PCR performed using samples collected by Direct Saliva Sample Collection method (DiSC).

|  |  | RT-PCR |  |  |
| --- | --- | --- | --- | --- |
|  |  | Positive | Negative | Total |
| TMA | Positive | 21 | 6 | 27 |
|  | Negative | 2 | 15 | 17 |
|  | Total | 23 | 21 | 44 |

TMA against RT-PCR
|  |  |
| --- | --- |
| positive percent agreement | 91% |
| negative percent agreement | 71% |
| Overall Agreement | 82% |

## Data Availability

All data produced in the present study are available upon reasonable request to the authors

## ICMJE Statement

Contributors YI was responsible for the organization and coordination of the trial. KA and KS were the chief investigators and responsible for the data analysis. KA, MN, TC, KS, YI and KT developed the trial design. All authors contributed to the writing of the final manuscript. All members of the DiSC Study Team contributed to the management or administration of the trial.

